# SARS-CoV-2 Omicron subvariants Spike recognition and neutralization elicited after the third dose of mRNA vaccine

**DOI:** 10.1101/2022.08.03.22278386

**Authors:** Alexandra Tauzin, Alexandre Nicolas, Shilei Ding, Mehdi Benlarbi, Halima Medjahed, Debashree Chatterjee, Katrina Dionne, Shang Yu Gong, Gabrielle Gendron-Lepage, Yuxia Bo, Josée Perreault, Guillaume Goyette, Laurie Gokool, Pascale Arlotto, Chantal Morrisseau, Cécile Tremblay, Valérie Martel-Laferrière, Gaston De Serres, Inès Levade, Daniel E. Kaufmann, Marceline Côté, Renée Bazin, Andrés Finzi

## Abstract

Several SARS-CoV-2 Omicron subvariants have recently emerged, becoming the dominant circulating strains in many countries. These variants contain a large number of mutations in their Spike glycoprotein, raising concerns about vaccine efficacy. In this study, we evaluate the ability of plasma from a cohort of individuals that received three doses of mRNA vaccine to recognize and neutralize these Omicron subvariant Spikes. We observed that BA.4/5 and BQ.1.1 Spikes are markedly less recognized and neutralized compared to the D614G and the other Omicron subvariant Spikes tested. Also, individuals who have been infected before or after vaccination present better humoral responses than SARS-CoV-2 naïve vaccinated individuals, thus indicating that hybrid immunity generates better humoral responses against these subvariants.

## INTRODUCTION

The SARS-CoV-2 Omicron variant BA.1 emerged at the end of 2021 and rapidly became the dominant circulating strain in the world ^1,2^. Since its emergence, several sublineages of Omicron rapidly replaced the BA.1 variant due to higher transmission rates. BA.2 became the dominant circulating strain in spring 2022 ^3,4^, and currently the BA.4 and BA.5 variants [sharing the same mutations in their Spike (S) glycoproteins, named BA.4/5 S in the manuscript], are the dominant circulating strains in several countries ^5–8^. BA.2.75, BA.4.6 and BQ.1.1 have emerged more recently and are spreading worldwide ^9,10^.

It was previously shown that poor humoral responses against BA.1 and BA.2 variants were observed after two doses of mRNA vaccine ^11–13^. We and other reported that an extended interval between the first two doses of mRNA vaccine led to strong humoral responses to several variants of concern (VOCs) including BA.1 and BA.2 after the second dose of mRNA vaccine ^14–16^. However, a third dose of mRNA vaccine led to an increase of humoral responses against these Omicron variants, regardless of the interval between doses ^11,13,16,17^. Previous studies also reported that breakthrough infection (BTI) in vaccinated people induced strong neutralizing Abs against VOCs, including BA.1 ^18,19^. However, recent studies have shown that BA.4/5, BA.2.75, BA.4.6 and BQ.1.1 appear to be more resistant than BA.1 and BA.2 to vaccination and monoclonal antibodies (Abs) ^20–26^.

In this study, we analyzed the ability of plasma from vaccinated individuals to recognize and neutralize pseudoviral particles bearing different Omicron subvariant Spikes four weeks (median [range]: 30 days [20–44 days]) and four months (median [range]: 121 days [92–135 days]) after the third dose of mRNA vaccine. This study was conducted in a cohort of individuals who received their first two doses with a 16-weeks extended interval (median [range]: 110 days [54– 146 days]) and their third dose seven months after the second dose (median [range]: 211 days [151-235 days]). The cohort included 15 naïve individuals who were never infected with SARS-

CoV-2, 15 previously infected (PI) individuals who were infected during the first wave of COVID-19 in early 2020 (before the advent of the alpha variant and other VOCs) and before vaccination, and 15 BTI individuals who were infected after vaccination. All BTI individuals were infected between mid-December 2021 and May 2022, when almost only Omicron variants (BA.1 and BA.2) were circulating in Quebec. Basic demographic characteristics of the cohorts and detailed vaccination time points are summarized in Table 1 and Figure 1A.

**Table 1.** Characteristics of the vaccinated SARS-CoV-2 cohorts.

|  |  | Entire cohort | Naïve | Breakthrough infection<sup>b</sup> | Previously infected |
| --- | --- | --- | --- | --- | --- |
| **Number** |  | 45 | 15 | 15 | 15 |
| **Age** |  | 51 (24-67) | 54 (24-67) | 43 (30-64) | 48 (29-65) |
| **Gender** | **Male (n)** | 17 | 4 | 5 | 8 |
|  | **Female (n)** | 28 | 11 | 10 | 7 |
| **Vaccine** | **1<sup>st</sup> dose** | Pfz=43 ; M=1 ; AZ=1 | Pfz=14 ; M=1 | Pfz=14 ; AZ=1 | Pfz=15 |
|  | **2<sup>nd</sup> dose** | Pfz=43 ; M=1 ; AZ=1 | Pfz=14 ; M=1 | Pfz=14 ; AZ=1 | Pfz=15 |
|  | **3<sup>rd</sup> dose** | Pfz=40 ; M=5 | Pfz=15 | Pfz=14 ; AZ=1 | Pfz=11 ; AZ=4 |
| **Days between symptom onset and the 1<sup>st</sup> dose<sup>a</sup>** |  | N/A | N/A | N/A | 288 (166-321) |
| **Days between the 1<sup>st</sup> and 2<sup>nd</sup> dose<sup>a</sup>** |  | 110 (54-146) | 109 (65-120) | 110 (54-113) | 112 (90-146) |
| **Days between the 2<sup>nd</sup> and 3<sup>rd</sup> dose<sup>a</sup>** |  | 211 (151-235) | 210 (184-227) | 215 (151-224) | 219 (187-235) |
| **Days between the 3<sup>rd</sup> dose and 4W** |  | 30 (20-44) | 32 (21-37) | 28 (20-38) | 33 (24-44) |
| **Days between the 3<sup>rd</sup> dose and 4M** |  | 121 (92-135) | 124 (105-135) | 121 (92-131) | 119 (111-127) |

<sup>a</sup> Values displayed are medians, with ranges in parentheses. Continuous variables were compared by using Kruskal- Wallis tests. $p < 0.05$ was considered statistically significant for all analyses. No statistical differences were found for any of the parameter tested between the different groups. Pfz= Pfizer/BioNtech BNT162b2, M= Moderna mRNA-1273, AZ= AstraZeneca ChAdOx1. <sup>b</sup> All Breakthrough infection individuals were infected between mid-December 2021 and May 2022, when almost only Omicron variants (BA.1 and BA.2) were circulating in Quebec. 6 BTI individuals were infected before the time point collected 4 weeks after the third dose and 9 BTI individuals were infected between the two time points.

**Figure 1.**
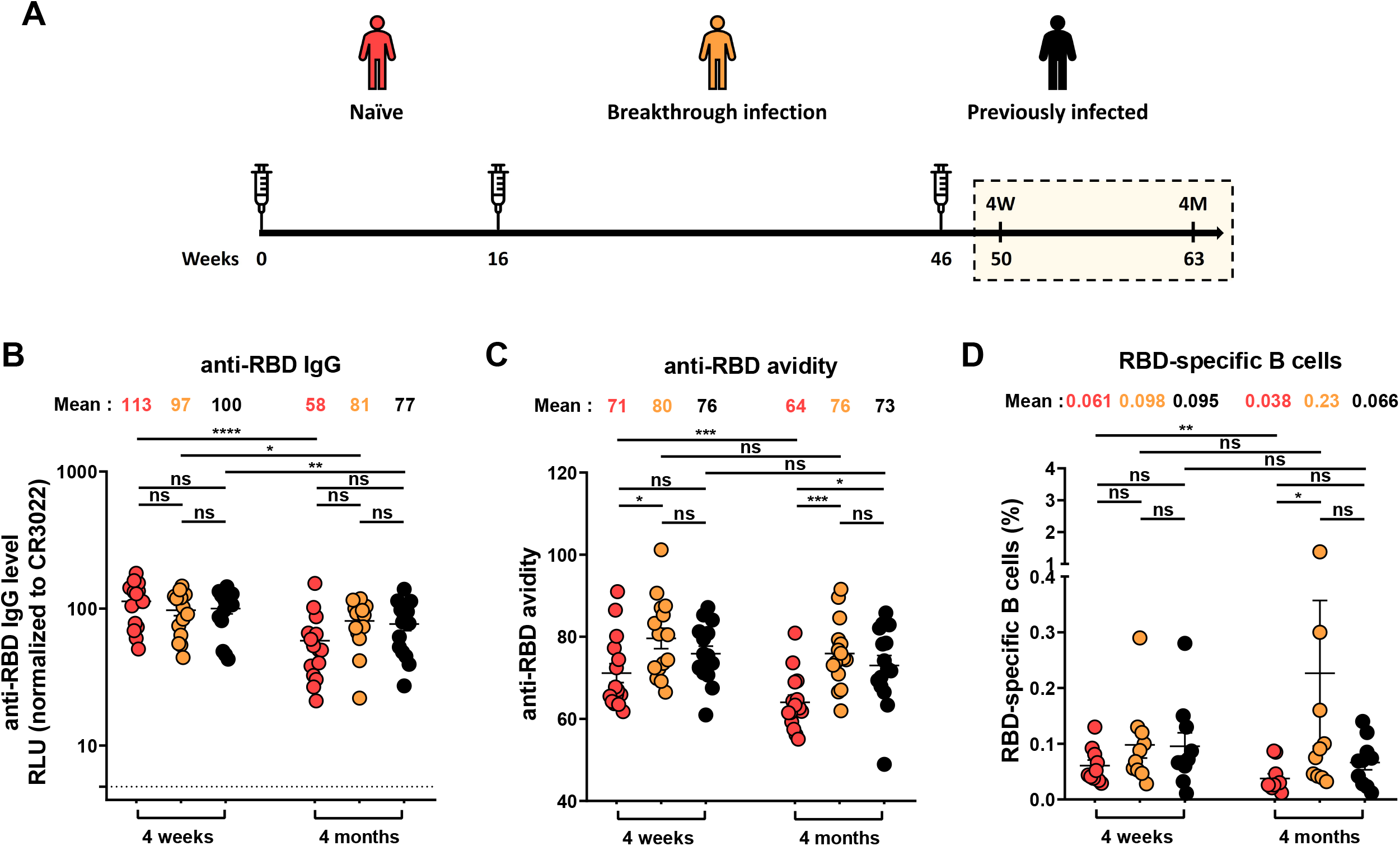
Anti-RBD IgG level, associated anti-RBD avidity and RBD-Specific B cell responses in plasma from naïve, BTI and PI individuals after the third dose of mRNA vaccine. (**A**) SARS-CoV-2 vaccine cohort design. The yellow box represents the period under study. (**B-C**) Indirect ELISAs were performed by incubating plasma samples from naïve, BTI or PI individuals collected 4 weeks or 4 months after the third dose of mRNA vaccine with recombinant SARS-CoV-2 RBD protein. Anti-RBD Ab binding was detected using HRP-conjugated anti-human IgG. (**B**) RLU values obtained were normalized to the signal obtained with the anti-RBD CR3022 mAb present in each plate. (**C**) The RBD avidity index corresponded to the value obtained with the stringent (8M urea) ELISA divided by that obtained without urea. (**D**) The frequencies of RBD+ B cells were measured by flow cytometry. (**B-D**) Plasma samples were grouped in two different time points (4 weeks and 4 months). Naïve, BTI and PI individuals are represented by red, yellow and black points respectively, undetectable measures are represented as white symbols, and limits of detection are plotted. Error bars indicate means ± SEM. (* P < 0.05; ** P < 0.01; *** P < 0.001; **** P < 0.0001; ns, non-significant). For all groups, n=15 (**B-C**) or n=10 (**D**).

## RESULTS

### RBD-specific IgG and associated avidity

We first measured the level of anti-RBD IgG four weeks and four months after the third dose of mRNA vaccine in naïve, BTI and PI individuals (Figure 1A) by a well described ELISA assay ^16,27–30^ (Figure 1B). Four weeks after the third dose, we did not observe significant differences in the level of IgG between the three groups. Four months after the third dose, we observed that the level of IgG significantly decreased in all groups, but to a higher extent in naïve individuals. No significant differences were observed between naïve, BTI and PI individuals four months after the boost. We also measured the avidity of anti-RBD IgG induced after the third dose of mRNA vaccine using a previously described assay ^28,31^. Four weeks after the third dose, we observed that naïve donors had IgG with lower avidity, although we only measured significant difference with BTI individuals (Figure 1C). Four months after the third dose, the avidity of these IgG slightly decreased for naïve individuals but remained stable for BTI and PI groups.

### RBD-specific B cell responses after the third dose of mRNA vaccine

We also monitored the SARS-CoV-2-specific B cells (identified as CD19+ CD20+) by flow cytometry, using two recombinant RBD protein probes labeled with two different fluorochromes (Alexa Fluor 594 and Alexa Fluor 488) (Figure S1A) ^30,32^. Four weeks after the third dose of mRNA vaccine, no significant differences in the level of circulating B cells were observed between the three groups (Figure 1D). Four months after the boost, this level significantly decreased for naïve donors but not for individuals with hybrid immunity (PI and BTI). For BTI individuals, we observed an increase, with some donors presenting a higher level of circulating RBD-specific B cells, probably due to recent infection.

### Recognition of SARS-CoV-2 Spike variants by plasma from vaccinated individuals

We next measured the ability of plasma to recognize the SARS-CoV-2 D614G and different Omicron subvariants S in vaccinated naïve, PI and BTI individuals four weeks and four months after the third dose of mRNA vaccine. Spike expression levels of the Spike variants were normalized to the signal obtained with the conformationally independent anti-S2 neutralizing CV3-25 antibody ^33–35^ that efficiently recognized all these Spikes despite their various mutations (Figure S1B, S2A-C). Four weeks after the third dose of mRNA vaccine, we observed that plasma from PI individuals recognized more efficiently the D614G S than naïve individuals (Figure 2A). We also observed that BTI individuals recognized the D614G S as efficiently as the PI individuals. Four months after the third dose, the level of recognition of the D614G S decreased for the three groups but with a more significant reduction in the naïve group. For the BA.2, BA.4/5 and BQ.1.1 S, naïve and BTI had the same level of recognition four weeks after the third dose, and this level was significantly lower than for PI individuals (Figure 2B-C, F). For BA.2.75 and BA.4.6 S, we only observed significant differences between naïve and PI individuals four weeks after the third dose (Figure 2D-E). Four months after the third dose, we observed a significant decrease of the recognition for naïve and PI individuals, with the exception of the BQ.1.1 S for which the level remained stable in the PI group (Figure 2A-F). For the BTI group, the level of recognition remained more stable and reached the same level than for the PI group for all tested Spikes. We also observed that the BA.4/5 and the BQ.1.1 S were always less recognized than the D614G and other Omicron subvariants S at both time points for all groups (Figure 2G-H).

**Figure 2.**
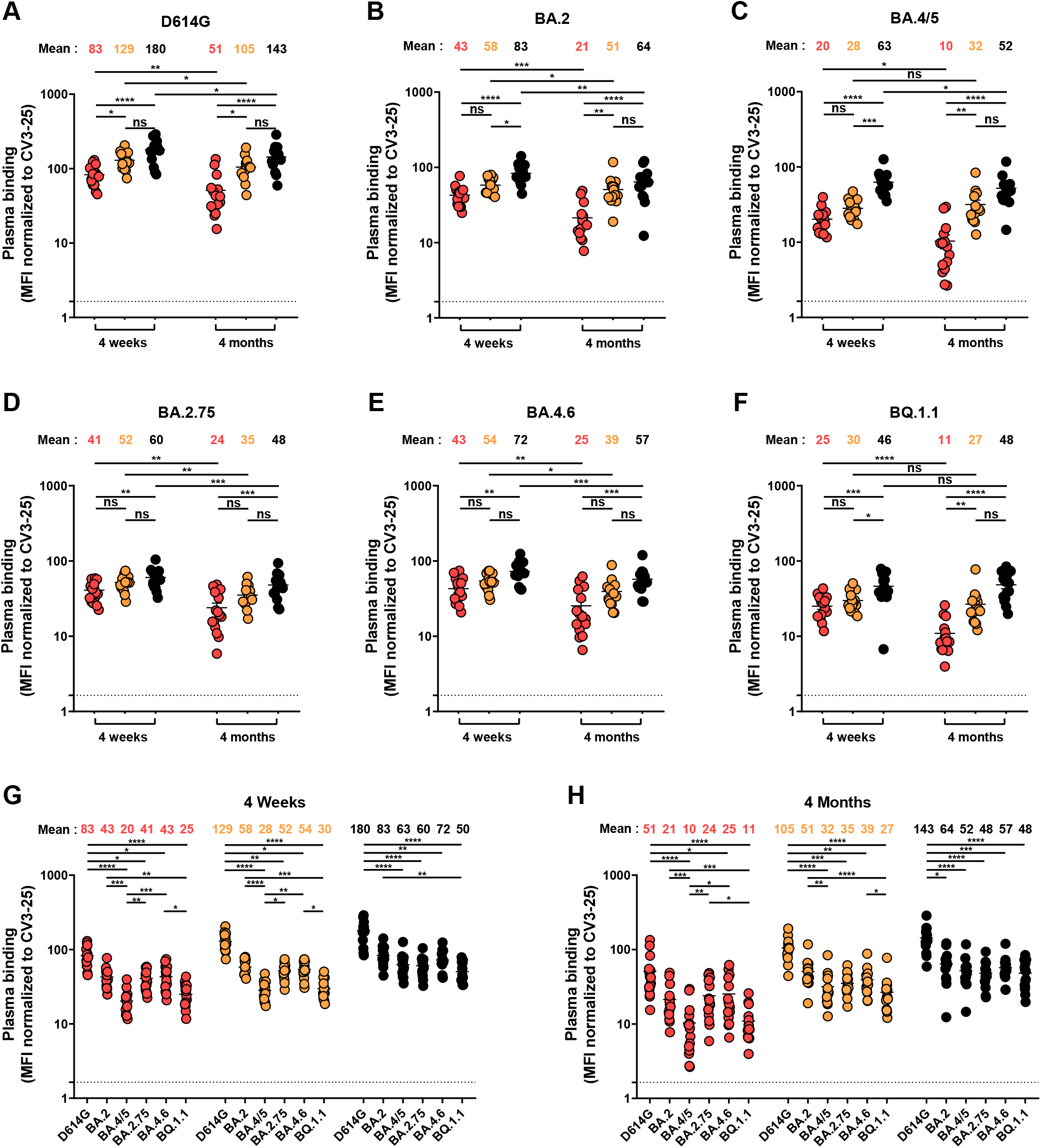
Recognition of SARS-CoV-2 Spike variants by plasma from naïve, BTI and PI individuals after the third dose of mRNA vaccine. (**A-G**) 293T cells were transfected with the indicated full-length S from different SARS-CoV-2 variants and stained with the CV3-25 mAb or with plasma from naïve, BTI or PI individuals collected 4 weeks or 4 months after the third dose of mRNA vaccine and analyzed by flow cytometry. The values represent the MFI normalized by CV3-25 Ab binding. (**A-F**) Plasma samples were grouped in two different time points (4 weeks and 4 months). (**G-H**) Binding of plasma collected at 4 weeks (**G**) and 4 months (**H**) post vaccination were measured. Naïve, BTI and PI individuals are represented by red, yellow and black points respectively, undetectable measures are represented as white symbols, and limits of detection are plotted. Error bars indicate means ± SEM. (* P < 0.05; ** P < 0.01; *** P < 0.001; **** P < 0.0001; ns, non-significant). For all groups, n=15.

### Neutralizing activity of the vaccine-elicited antibodies

We also evaluated the neutralizing activity against pseudoviral particles bearing these Spikes in the three groups. Of note, all Spikes were incorporated into pseudoviral particles to similar extents (Figure S2C) and had similar levels of infectivity in our assay (Figure S2D). In agreement with the pattern of S recognition, PI individuals neutralized more efficiently all the S variants tested than naïve individuals four weeks after the third dose (Figure 3A-F). For the BTI group, the level of neutralizing Abs was intermediate between the two other groups. Four months after the third dose, we did not observe significant differences between PI and BTI individuals. In contrast, the naïve group neutralized less efficiently the D614G, and Omicron subvariants S (Figure 3A-F). Four weeks after the third dose, no significant difference in the level of neutralization was measured between the D614G and BA.2 S for the three groups (Figure 2G). In contrast, the other Omicron variants S were more resistant to neutralization than the D614G S in all groups. Four months after the third dose, weak or no neutralizing activity against Omicron subvariants S was detected in most naïve individuals (Figure 3B-F, H). For BTI and PI individuals, although neutralizing activity was higher than in naïve individuals, the BA.4/5, BA.2.75, BA.4.6 and BQ.1.1 S were also significantly less neutralized than the D614G and, in some instances, BA.2 S (Figure 3B-F, H).

**Figure 3.**
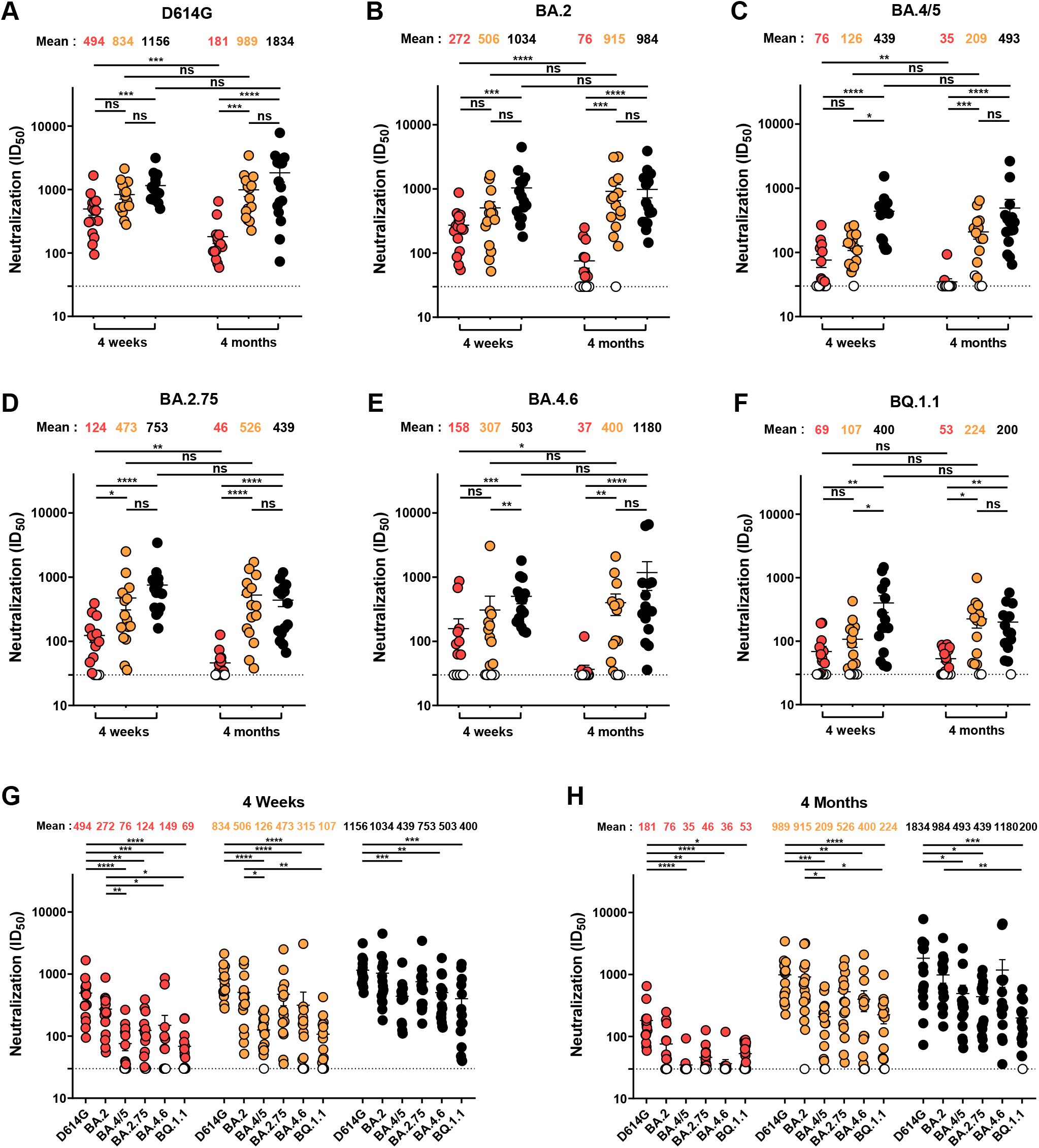
Neutralization activity of SARS-CoV-2 Spike variants by plasma from naïve, BTI and PI individuals after the third dose of mRNA vaccine. (**A-G**) Neutralization activity was measured by incubating pseudoviruses bearing SARS-CoV-2 S glycoproteins, with serial dilutions of plasma for 1 h at 37°C before infecting 293T-ACE2 cells. Neutralization half maximal inhibitory serum dilution (ID_50_) values were determined using a normalized non-linear regression using GraphPad Prism software. (**A-F**) Plasma samples were grouped in two different time points (4 weeks and 4 months). (**G-H**) Neutralization activity of plasma collected at 4 weeks (**G**) and 4 months (**H**) post vaccination were measured. Naïve, BTI and PI individuals are represented by red, yellow and black points respectively, undetectable measures are represented as white symbols, and limits of detection are plotted. Error bars indicate means ± SEM. (* P < 0.05; ** P < 0.01; *** P < 0.001; **** P < 0.0001; ns, non-significant). For all groups, n=15.

## DISCUSSION

More than two years after its emergence, and although an important proportion of the world population has received several doses of vaccine, the SARS-CoV-2 variants continue to circulate globally. In recent months, new sub-variants of Omicron emerged, carrying increasing numbers of mutations making them more transmissible and resistant to vaccination and monoclonal antibodies treatment ^8,17,20–22,25^. In agreement with this, we observed that the BA.4/5, BA.2.75, BA.4.6 and BQ.1.1 S were less efficiently recognized and neutralized than the D614G and the BA.2 S by plasma from individuals who received three doses of mRNA vaccine.

Several studies reported that poor neutralizing activity against VOCs was observed after two doses of mRNA vaccine, but a third dose strongly improved this response ^11,16,36^. However, when the second dose of vaccine was administered with an extended 16-weeks interval, higher humoral responses against VOCs (including BA.1 and BA.2) were observed after the second dose of vaccine ^14^, that were not increased by a booster dose ^16^. Therefore, there is no evidence that additional doses of the original SARS-CoV-2 vaccines after the third dose will result in increased responses against VOCs.

The Omicron variants spread more easily in vaccinated individuals than pre-Omicron variants ^37,38^. Interestingly, it was recently shown that previous infection with an Omicron variant prevents reinfection more efficiently than previous infection with a pre-Omicron variant ^39,40^, thus suggesting that new vaccines based on Omicron variants may generate humoral responses more likely to control Omicron sub-variants.

It was previously shown that hybrid immunity due to SARS-CoV-2 infection followed by vaccination confers stronger immune responses than vaccination alone ^16,32,40,41^. Accordingly, here we observed that individuals with BTI had the same level of S recognition and neutralization than individuals previously infected supporting the concept that hybrid protection is similar whatever the order of infection and vaccination. However, the durability of these responses remains unknown.

In conclusion, virus recognition and neutralizing activity induced by current mRNA vaccine are low against Omicron subvariants, rapidly decline over 4 months in naïve individuals, and will likely decrease further with future SARS-CoV-2 evolution. There is a need to rapidly develop new generations of vaccines that will elicit broader and less labile protection.

## LIMITATIONS OF THE STUDY

One of the limitations of the study is that for most BTI individuals, we do not have the exact day of infection and by which variant. We can only confirm whether they were infected before the first timepoint studied or between the 2 timepoints. Furthermore, it is very likely that some PI individuals were exposed a second time. However, in our study, no case of infection was confirmed by PCR in PI group, and since they were already infected a first time, we cannot conclude that a positive anti-N corresponds to a new infection or to the first. Finally, while we did not observe major differences in infectivity with our pseudoviral particles, it is possible that differences in infectivity and replication exist when using authentic live viruses. For this reason we only report on plasma neutralization profiles, which were shown to be similar between pseudoviral particles and authentic viruses and have been largely used by the field to inform on neutralizing responses elicited by natural infection and vaccination ^42–46^.

## Data Availability

All data produced in the present work are contained in the manuscript

## ACKNOWLEDGMENTS

The authors are grateful to the individuals who participated in this study. The authors thank the CRCHUM BSL3 and Flow Cytometry Platforms for technical assistance. This work was supported by le Ministère de l’Économie et de l’Innovation du Québec, Programme de soutien aux organismes de recherche et d’innovation to A.F. and by the Fondation du CHUM. This work was also supported by a CIHR foundation grant #352417, by a CIHR operating Pandemic and Health Emergencies Research grant #177958, by an Exceptional Fund COVID-19 from the Canada Foundation for Innovation (CFI) #41027 to A.F. and by a FRQS Merit Research Scholar award (# 268471) to D.E.K. Work on variants presented was also supported by the Sentinelle COVID Quebec network led by the LSPQ in collaboration with Fonds de Recherche du Québec Santé (FRQS) to A.F. This work was also partially supported by a CIHR COVID-19 rapid response grant (OV3 170632) and CIHR stream 1 SARS-CoV-2 Variant Research to M.C. A.F. is the recipient of Canada Research Chair on Retroviral Entry no. RCHS0235 950-232424. M.C is a Tier II Canada Research Chair in Molecular Virology and Antiviral Therapeutics (950-232840).

C.T. is the Pfizer/Université de Montréal Chair on HIV translational research. A.T. was supported by MITACS Accélération postdoctoral fellowship. M.B. was the recipient of a CIHR master’s scholarship award. The funders had no role in study design, data collection and analysis, decision to publish, or preparation of the manuscript. We declare no competing interests.

## AUTHOR CONTRIBUTIONS

A.T. and A.F. conceived the study. A.T., A.N., S.D., D.C., M.B., K.D., S.Y.G., G.G.L., H.M., G.G., J.P., Y.B., and A.F. performed, analyzed, and interpreted the experiments. A.T. performed statistical analysis. G.G.L., H.M., G.G., M.C., and A.F. contributed unique reagents. L.G., P.A., C.M., C.T., and V.M.-L. collected and provided clinical samples. R.B., G.D.S., D.E.K. and I.L. provided scientific input related to VOCs and vaccine efficacy. A.T. and A.F. wrote the manuscript with inputs from others. Every author has read, edited, and approved the final manuscript.

## STAR METHODS

### RESOURCE AVAILABILITY

#### Lead contact

Further information and requests for resources and reagents should be directed to and will be fulfilled by the lead contact, Andrés Finzi.

#### Materials availability

All unique reagents generated during this study are available from the Lead contact without restriction.

#### Data and code availability

- All data reported in this paper will be shared by the lead contact upon request.
- This paper does not report original code.
- Any additional information required to reanalyze the data reported in this paper is available from the lead contact upon request.

### EXPERIMENTAL MODEL AND SUBJECT DETAILS

#### Ethics Statement

All work was conducted in accordance with the Declaration of Helsinki in terms of informed consent and approval by an appropriate institutional board. Blood samples were obtained from donors who consented to participate in this research project at CHUM (19.381). Plasmas and PBMCs were isolated by centrifugation and Ficoll gradient, and samples stored at -80°C and in liquid nitrogen respectively, until use.

#### Human subjects

The study was conducted in 15 SARS-CoV-2 naïve individuals (4 males and 11 females; age range: 24-67 years), 15 SARS-CoV-2 breakthrough infection individuals (5 males and 10 females; age range: 30-64 years) infected after the second or third dose of mRNA vaccine (6 BTI were infected before the time point collected 4 weeks after the third dose and 9 individuals were infected between the two time points), and 15 SARS-CoV-2 previously infected individuals (8 males and 7 females; age range: 29-65 years) infected before vaccination during the first wave of COVID-19 in march-may 2020. This information is presented in table 1. No specific criteria such as number of patients (sample size), gender, clinical or demographic were used for inclusion, beyond PCR confirmed SARS-CoV-2 infection in adults before vaccination for PI group, PCR confirmed SARS-CoV-2 infection or anti-N positive in adults after vaccination for BTI group and no detection of Abs recognizing the N protein for naïve individuals.

#### Plasma and antibodies

Plasmas were isolated by centrifugation with Ficoll gradient, heat-inactivated for 1 hour at 56°C and stored at -80°C until use in subsequent experiments. Healthy donor’s plasmas, collected before the pandemic, were used as negative controls, and used to calculate the seropositivity threshold in our ELISAs and flow cytometry assays (data not shown). The RBD-specific monoclonal antibody CR3022 was used as a positive control in ELISA assays, and the conformationally independent S2-specific monoclonal antibody CV3-25 was used as a positive control and to normalize Spike expression in our flow cytometry assays, as described ^34,47–49^. Horseradish peroxidase (HRP)-conjugated Abs able to detect the Fc region of human IgG (Invitrogen) was used as secondary Abs to detect Ab binding in ELISA experiments. Alexa Fluor-647-conjugated goat anti-human Abs able to detect all Ig isotypes (anti-human IgM+IgG+IgA; Jackson ImmunoResearch Laboratories) were used as secondary Abs to detect plasma binding in flow cytometry experiments.

#### Cell lines

293T human embryonic kidney cells (obtained from ATCC) and Cf2.Th cells (a kind gift from Joseph Sodroski, Dana Farber Cancer Institute (DFCI), Boston, MA, USA) were maintained at 37°C under 5% CO_2_ in Dulbecco’s modified Eagle’s medium (DMEM) (Wisent) containing 5% fetal bovine serum (FBS) (VWR) and 100 μg/ml of penicillin-streptomycin (Wisent). 293T-ACE2 cell line was previously reported ^29^.

### METHOD DETAILS

#### Plasmids

The plasmids encoding the SARS-CoV-2 Spike variants were previously described ^16,48,50^. The plasmids encoding the BA.4/5, BA.2.75, BA.4.6 and BQ.1.1 Spike was generated by overlapping PCR using the BA.2 SARS-CoV-2 Spike gene as a template and cloned in pCAGGS. All constructs were verified by Sanger sequencing. Spike variant sequences are outlined in Figure S2A. The pNL4.3 R-E-Luc was obtained from the NIH AIDS Reagent Program. The vesicular stomatitis virus G (VSV-G)-encoding plasmid (pSVCMV-IN-VSV-G) was previously described ^29^.

#### Protein expression and purification

FreeStyle 293F cells (Invitrogen) were grown in FreeStyle 293F medium (Invitrogen) to a density of 1×10^6^ cells/mL at 37°C with 8% CO_2_ with regular agitation (150 rpm). Cells were transfected with a plasmid coding for SARS-CoV-2 S WT RBD ^50^ using ExpiFectamine 293 transfection reagent, as directed by the manufacturer (Invitrogen). One week later, cells were pelleted and discarded. Supernatants were filtered using a 0.22 μm filter (Thermo Fisher Scientific). The recombinant RBD proteins were purified by nickel affinity columns, as directed by the manufacturer (Invitrogen). The RBD preparations were dialyzed against phosphate-buffered saline (PBS) and stored in aliquots at -80°C until further use. To assess purity, recombinant proteins were loaded on SDS-PAGE gels and stained with Coomassie Blue.

#### Enzyme-linked immunosorbent assay (ELISA) and RBD avidity index

The SARS-CoV-2 WT RBD ELISA assay used was previously described ^29,50^. Briefly, recombinant SARS-CoV-2 WT RBD proteins (2.5 mg/mL), or bovine serum albumin (BSA) (2.5 mg/mL) as a negative control, were prepared in PBS and were adsorbed to plates (MaxiSorp Nunc) overnight at 4°C. Coated wells were subsequently blocked with blocking buffer (Tris-buffered saline [TBS] containing 0.1% Tween20 and 2% BSA) for 1h at room temperature. Wells were then washed four times with washing buffer (Tris-buffered saline [TBS] containing 0.1% Tween20). CR3022 mAb (50 ng/mL) or a 1/500 dilution of plasma were prepared in a diluted solution of blocking buffer (0.1% BSA) and incubated with the RBD-coated wells for 90 min at room temperature. Plates were washed four times with washing buffer followed by incubation with secondary Abs (diluted in a diluted solution of blocking buffer (0.4% BSA)) for 1h at room temperature, followed by four washes. To calculate the RBD-avidity index, we performed in parallel a stringent ELISA, where the plates were washed with a chaotropic agent, 8M of urea, added of the washing buffer. This assay was previously described ^31^. HRP enzyme activity was determined after the addition of a 1:1 mix of Western Lightning oxidizing and luminol reagents (Perkin Elmer Life Sciences). Light emission was measured with a LB942 TriStar luminometer (Berthold Technologies). Signal obtained with BSA was subtracted for each plasma and was then normalized to the signal obtained with CR3022 present in each plate. The seropositivity threshold was established using the following formula: mean of pre-pandemic SARS-CoV-2 negative plasma + (3 standard deviation of the mean of pre-pandemic SARS-CoV-2 negative plasma).

#### SARS-CoV-2-specific B cells characterization

To detect SARS-CoV-2-specific B cells, we conjugated recombinant RBD proteins with Alexa Fluor 488 or Alexa Fluor 594 (Thermo Fisher Scientific) according to the manufacturer’s protocol. 2×10^6^ frozen PBMCs from SARS-CoV-2 naïve, BTI and PI donors were prepared at a final concentration of 4×10^6^ cells/mL in RPMI 1640 medium (GIBCO) supplemented with 10% of fetal bovine serum (Seradigm), Penicillin/Streptomycin (GIBCO) and HEPES (GIBCO). After a rest of 2h at 37°C and 5% CO_2_, cells were stained using Aquavivid viability marker (GIBCO) in DPBS (GIBCO) at 4°C for 20 min. The detection of SARS-CoV-2-antigen specific B cells was done by adding the RBD probes to the antibody cocktail listed in Table S1. Staining was performed at 4°C for 30 min and cells were fixed using 2% paraformaldehyde at 4°C for 15 min. Stained PBMC samples were acquired on Symphony cytometer (BD Biosciences) and analyzed using FlowJo v10.8.0 software and the gating strategy presented in Figure S1A.

#### Cell surface staining and flow cytometry analysis

293T were transfected with full-length SARS-CoV-2 Spikes and a green fluorescent protein (GFP) expressor (pIRES2-eGFP; Clontech) using the calcium-phosphate method. Two days post-transfection, Spike-expressing 293T cells were stained with the CV3-25 Ab (5 μg/mL) as control or plasma from naïve, BTI or PI individuals (1:250 dilution) for 45 min at 37°C. AlexaFluor-647-conjugated goat anti-human IgG (1/1000 dilution) were used as secondary Abs. The percentage of Spike-expressing cells (GFP + cells) was determined by gating the living cell population based on viability dye staining (Aqua Vivid, Invitrogen). Samples were acquired on a LSR II cytometer (BD Biosciences), and data analysis was performed using FlowJo v10.7.1 (Tree Star) using the gating strategy presented in Figure S1B. The conformationally-independent anti-S2 antibody CV3-25 was used to normalize Spike expression, as reported ^33–35,48^. CV3-25 was shown to be effective against all Spike variants (Figure S2B-C). The Median Fluorescence intensities (MFI) obtained with plasma were normalized to the MFI obtained with CV3-25 and presented as percentage of CV3-25 binding.

#### Pseudoviral infectivity

293T cells were transfected with the lentiviral vector pNL4.3 R-E-Luc (NIH AIDS Reagent Program) and plasmid encoding for the indicated S glycoprotein (D614G, BA.2, BA.4/5, BA.2.75, BA.4.6 or BQ.1.1) at a ratio of 10:1. Two days post-transfection, cell supernatants were harvested and stored at -80°C until use. The RT activity was evaluated by measure of the incorporation of [*methyl*-3H]TTP into cDNA of a poly(rA) template in the presence of virion-associated RT and oligo(dT). Normalized amount of RT activity pseudoviral particles were added to 293T-ACE2 target cells for 48 h at 37°C. Then, cells were lysed by the addition of 30 μL of passive lysis buffer (Promega) followed by one freeze-thaw cycle. An LB942 TriStar luminometer (Berthold Technologies) was used to measure the luciferase activity of each well after the addition of 100 μL of luciferin buffer (15mM MgSO4, 15mM KPO4 [pH 7.8], 1mM ATP, and 1mM dithiothreitol) and 50 μL of 1mM d-luciferin potassium salt (Thermo Fisher Scientific). RLU values obtained were normalized to D614G.

#### Virus neutralization assay

To produce SARS-CoV-2 pseudoviruses, 293T cells were transfected with the lentiviral vector pNL4.3 R-E– Luc (NIH AIDS Reagent Program) and a plasmid encoding for the indicated S glycoprotein (D614G, BA.2, BA.4/5, BA.2.75, BA.4.6 or BQ.1.1) at a ratio of 10:1. Two days post-transfection, cell supernatants were harvested and stored at –80°C until use. For the neutralization assay, 293T-ACE2 target cells were seeded at a density of 1×10^4^ cells/well in 96-well luminometer-compatible tissue culture plates (PerkinElmer) 24h before infection. Pseudoviral particles were incubated with several plasma dilutions (1/50; 1/250; 1/1250; 1/6250; 1/31250) for 1h at 37°C and were then added to the target cells followed by incubation for 48h at 37°C. Cells were lysed by the addition of 30 μL of passive lysis buffer (Promega) followed by one freeze-thaw cycle. An LB942 TriStar luminometer (Berthold Technologies) was used to measure the luciferase activity of each well after the addition of 100 μL of luciferin buffer (15mM MgSO_4_, 15mM KH_2_PO_4_ [pH 7.8], 1mM ATP, and 1mM dithiothreitol) and 50 μL of 1mM d-luciferin potassium salt (Prolume). The neutralization half-maximal inhibitory dilution (ID_50_) represents the plasma dilution to inhibit 50% of the infection of 293T-ACE2 cells by pseudoviruses.

#### Virus Capture Assay

The assay was previously described ^51^. Briefly, pseudoviral particles were produced by transfecting 2×10^6^ 293T cells with pNL4.3 R-E-Luc (3.5 μg), pSVCMV-IN-VSV-G (1 μg) and plasmids encoding for SARS-CoV-2 Spike glycoproteins (3.5 μg) using the standard calcium phosphate protocol. 48 hours later, virion-containing supernatants were collected. White MaxiSorp ELISA plates (Thermo Fisher Scientific, Waltham, MA, USA.) were plated with the CV3-25 mAb at 0.05 μg per well overnight at 4°C. Unbound antibodies were removed by washing the plates twice with PBS. Plates were subsequently blocked with 3% BSA in PBS for 1 h at room temperature. After the washes, 200 μL of virus-containing supernatant was added to the wells. Viral capture by the Ab was visualized by adding 1×10^4^ SARS-CoV-2-resistant Cf2Th cells in full DMEM medium per well. Forty-eight hours post-infection, cells were lysed by the addition of 30 μL of passive lysis buffer (Promega, Madison, WI, USA.) and three freeze-thaw cycles. An LB941 TriStar luminometer (Berthold Technologies) was used to measure the luciferase activity of each well after the addition of 100 μL of luciferin buffer (15 mM MgSO4, 15 mM KH2PO4 (pH 7.8), 1 mM ATP, and 1 mM dithiothreitol) and 50 μL of 1 mM D-luciferin potassium salt (Prolume, Randolph, VT, USA.).

### QUANTIFICATION AND STATISTICAL ANALYSIS

#### Statistical analysis

Symbols represent biologically independent samples from SARS-CoV-2 naïve, BTI or PI individuals. Statistics were analyzed using GraphPad Prism version 8.0.1 (GraphPad, San Diego, CA). Every dataset was tested for statistical normality and this information was used to apply the appropriate (parametric or nonparametric) statistical test. p values < 0.05 were considered significant; significance values are indicated as ^*^P<0.05, ^**^P<0.01, ^***^P<0.001, ^***^P<0.0001, ns, non-significant.

## SUPPLEMENTAL INFORMATION

Supplemental information can be found online at …

---

**Table S1.** Flow cytometry antibody staining panel for B cells characterization. Related to Figure 1 and the STAR Methods section.

| **Marker-Fluorophore** | **Clone** | **Vendor** | **Catalog #** |
| --- | --- | --- | --- |
| CD3 – BV480 | UCHT1 | BD Biosciences | 566105 |
| CD14 – BV480 | M5E2 | BD Biosciences | 746304 |
| CD16 – BV480 | 3G8 | BD Biosciences | 566108 |
| CD19 – BV650 | SJ25C1 | Biolegend | 363026 |
| CD20 – BV711 | 2H7 | Biolegend | 302342 |
| CD21 – BV786 | B-LY4 | BD Biosciences | 740969 |
| CD24 – BUV805 | ML5 | BD Biosciences | 742010 |
| CD27 – APC-R700 | M-T271 | BD Biosciences | 565116 |
| CD38 – BB790 | HIT2 | BD Biosciences | CUSTOM |
| CD56 – BV480 | NCAM16.2 | BD Biosciences | 566124 |
| CD138 – BUV661 | MI15 | BD Biosciences | 749873 |
| CCR10 – BUV395 | 1B5 | BD Biosciences | 565322 |
| HLA-DR – BB700 | G46-6 | BD Biosciences | 566480 |
| IgA - PE | IS11-8E10 | Miltenyi Biotec | 130-113-476 |
| IgD – BUV563 | IA6-2 | BD Biosciences | 741394 |
| IgG – BV421 | G18-147 | BD Biosciences | 562581 |
| IgM – BUV737 | UCH-B1 | BD Biosciences | 748928 |
| LIVE/DEAD Fixable dead cell | N/A | Thermo Fisher Scientific | L34960 |

**Figure S1.**
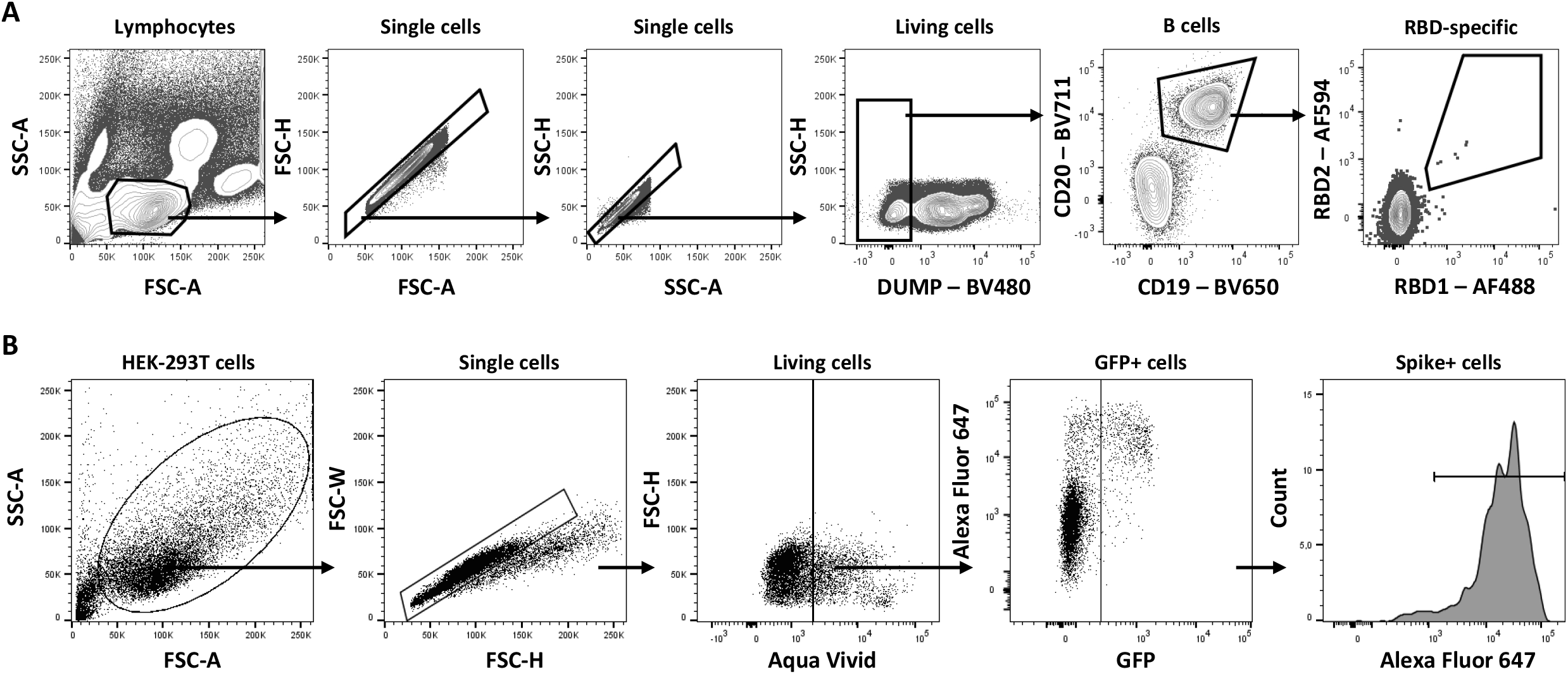
Gating strategy for SARS-CoV-2-specific B cell characterization and SARS-CoV-2 Spikes recognition, Related to Figures 1 and 2. **(A)** Representative flow cytometry gates to identify RBD-specific B cells. (**B**) Representative flow cytometry gates to identify the level of recognition of the Spikes. 293T cells were transfected with the full-length Spike from different SARS-CoV-2 variants (D614G, BA.2, BA.4/5, BA.2.75, BA.4.6 and BQ.1.1), stained with plasma or the CV3-25 mAb and analyzed by flow cytometry. Cells expressing the Spike were identified according to cell morphology by light-scatter parameters (first column) and excluding doublets cells (second column). Cells were then gated on living cells (excluding the dead cells labeled with Aqua vivid; third column). Finally, the percentage of GFP+ cells was used to measure the median of fluorescence of Alexa Fluor 647+ cells (last column).

**Figure S2.**
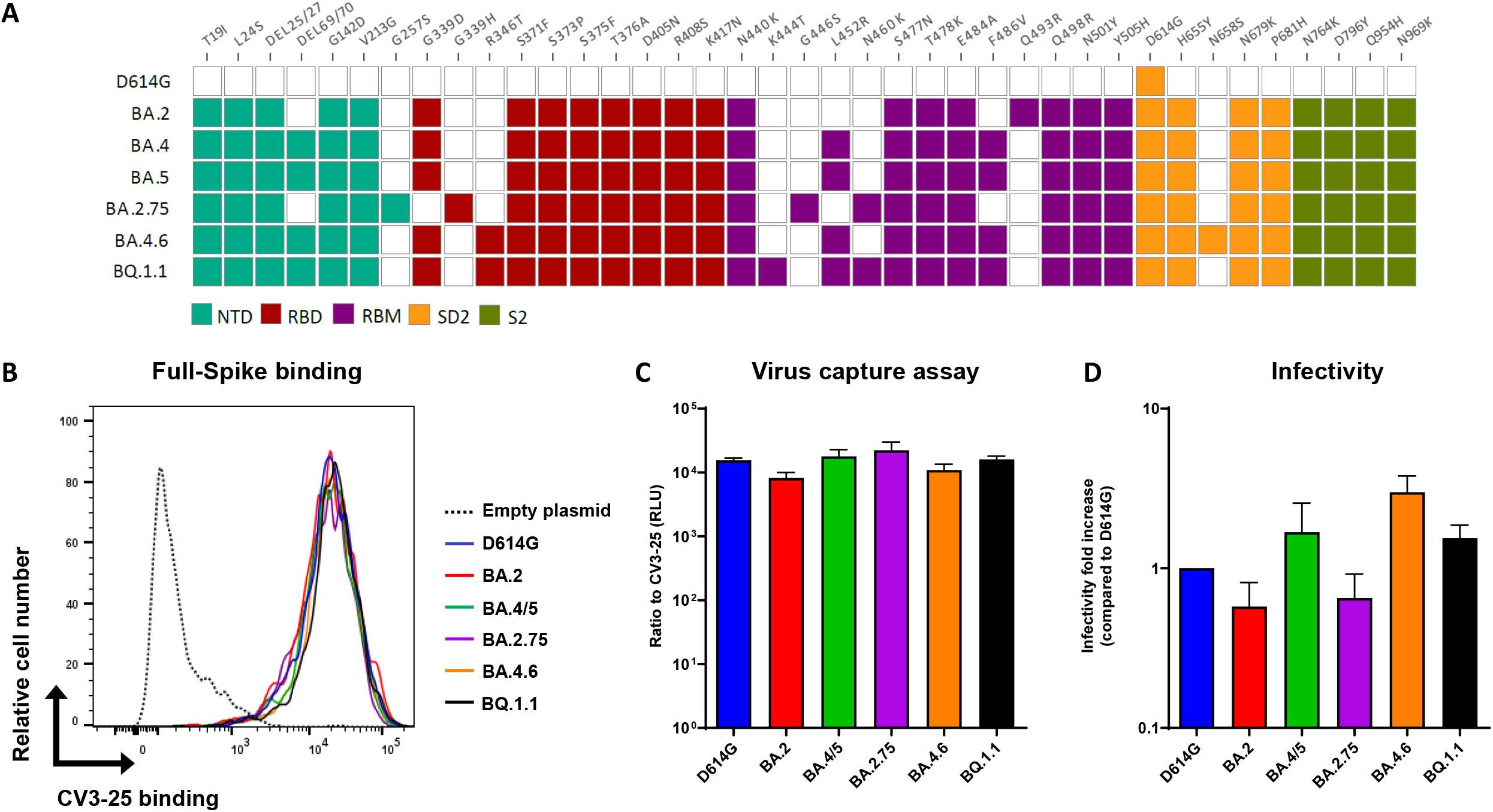
Recognition and infectivity of different Omicron subvariant Spikes, Related to Figures 2 and 3. **(A)** Mutations and deletions in the Spike amino acid sequence from D614G and Omicron subvariants compared to the ancestral strain. NTD: N-terminal domain; RBD: receptor-binding domain; RBM : receptor-binding motif; SD2: subdomain 2; S2: subunit 2. (**B**) 293T cells were transfected with the full-length Spike from different SARS-CoV-2 variants (D614G, BA.2, BA.4/5, BA.2.75, BA.4.6 and BQ.1.1), stained with the CV3-25 mAb and analyzed by flow cytometry. (**C**) VSV-G-pseudotyped viral particles expressing the indicated SARS-CoV-2 S glycoprotein were added to plates coated with CV3-25. Free virions were washed away, and Cf2Th cells were added to the wells. After 48 hours, cells were lysed, and the luciferase activity was measured. Luciferase signals were normalized to those obtained with the CV3-25 mAb. (**D**) Pseudoviral particles bearing the indicated SARS-CoV-2 S were used to infect 293T-ACE2 cells for 2 days at 37°C. RLU values obtained were normalized to D614G. (**C-D**) The experiments were repeated three times. Error bars indicate means ± SEM.

## REFERENCES

1. Viana, R., Moyo, S., Amoako, D.G., Tegally, H., Scheepers, C., Althaus, C.L., Anyaneji, U.J., Bester, P.A., Boni, M.F., Chand, M., et al. (2022). Rapid epidemic expansion of the SARS-CoV-2 Omicron variant in southern Africa. Nature, 1–10. 10.1038/s41586-022-04411-y.

2. WHO Update on Omicron, https://www.who.int/news/item/28-11-2021-update-on-omicron. https://www.who.int/news/item/28-11-2021-update-on-omicron.

3. CDC (2022). COVID Data Tracker Weekly Review, https://www.cdc.gov/coronavirus/2019-ncov/covid-data/covidview/past-reports/04222022.html. Cent. Dis. Control Prev. https://www.cdc.gov/coronavirus/2019-ncov/covid-data/covidview/past-reports/04222022.html.

4. Elliott, P., Eales, O., Steyn, N., Tang, D., Bodinier, B., Wang, H., Elliott, J., Whitaker, M., Atchison, C., Diggle, P.J., et al. (2022). Twin peaks: The Omicron SARS-CoV-2 BA.1 and BA.2 epidemics in England. Science 376, eabq4411. 10.1126/science.abq4411.

5. Mohapatra, R.K., Kandi, V., Sarangi, A.K., Verma, S., Tuli, H.S., Chakraborty, S., Chakraborty, C., and Dhama, K. (2022). The recently emerged BA.4 and BA.5 lineages of Omicron and their global health concerns amid the ongoing wave of COVID-19 pandemic – Correspondence. Int. J. Surg. Lond. Engl. 103, 106698. 10.1016/j.ijsu.2022.106698.

6. PHO (2022). Public Health Ontario: SARS-CoV-2 Omicron Variant Sub-Lineages BA.4 and BA.5: Evidence and Risk Assessment, chrome-extension://efaidnbmnnnibpcajpcglclefindmkaj/ https://www.publichealthontario.ca/-/media/Documents/nCoV/voc/2022/07/evidence-brief-ba4-ba5-risk-assessment-jul-8.pdf?sc_lang=en.

7. Tegally, H., Moir, M., Everatt, J., Giovanetti, M., Scheepers, C., Wilkinson, E., Subramoney, K., Makatini, Z., Moyo, S., Amoako, D.G., et al. (2022). Emergence of SARS-CoV-2 Omicron lineages BA.4 and BA.5 in South Africa. Nat. Med., 1–1. 10.1038/s41591-022-01911-2.

8. Yamasoba, D., Kosugi, Y., Kimura, I., Fujita, S., Uriu, K., Ito, J., and Sato, K. (2022). Neutralisation sensitivity of SARS-CoV-2 omicron subvariants to therapeutic monoclonal antibodies. Lancet Infect. Dis. 22, 942–943. 10.1016/S1473-3099(22)00365-6.

9. WHO (2022). Tracking SARS-CoV-2 variants, https://www.who.int/en/activities/tracking-SARS-CoV-2-variants/. https://www.who.int/health-topics/typhoid/tracking-SARS-CoV-2-variants.

10. IDSA (2022). SARS-CoV-2 Variants, https://www.idsociety.org/covid-19-real-time-learning-network/emerging-variants/emerging-covid-19-variants/. https://www.idsociety.org/covid-19-real-time-learning-network/emerging-variants/emerging-covid-19-variants/.

11. Muik, A., Lui, B.G., Wallisch, A.-K., Bacher, M., Mühl, J., Reinholz, J., Ozhelvaci, O., Beckmann, N., Güimil Garcia, R. de la C., Poran, A., et al. (2022). Neutralization of SARS-CoV-2 Omicron by BNT162b2 mRNA vaccine–elicited human sera. Science 375, 678–680. 10.1126/science.abn7591.

12. Nemet, I., Kliker, L., Lustig, Y., Zuckerman, N., Erster, O., Cohen, C., Kreiss, Y., Alroy-Preis, S., Regev-Yochay, G., Mendelson, E., et al. (2022). Third BNT162b2 Vaccination Neutralization of SARS-CoV-2 Omicron Infection. N. Engl. J. Med. 386, 492–494. 10.1056/NEJMc2119358.

13. Yu, J., Collier, A.Y., Rowe, M., Mardas, F., Ventura, J.D., Wan, H., Miller, J., Powers, O., Chung, B., Siamatu, M., et al. (2022). Neutralization of the SARS-CoV-2 Omicron BA.1 and BA.2 Variants. N. Engl. J. Med. 386, 1579–1580. 10.1056/NEJMc2201849.

14. Chatterjee, D., Tauzin, A., Marchitto, L., Gong, S.Y., Boutin, M., Bourassa, C., Beaudoin-Bussières, G., Bo, Y., Ding, S., Laumaea, A., et al. (2022). SARS-CoV-2 Omicron Spike recognition by plasma from individuals receiving BNT162b2 mRNA vaccination with a 16-week interval between doses. Cell Rep., 110429. 10.1016/j.celrep.2022.110429.

15. Payne, R.P., Longet, S., Austin, J.A., Skelly, D.T., Dejnirattisai, W., Adele, S., Meardon, N., Faustini, S., Al-Taei, S., Moore, S.C., et al. (2021). Immunogenicity of standard and extended dosing intervals of BNT162b2 mRNA vaccine. Cell 184, 5699-5714.e11. 10.1016/j.cell.2021.10.011.

16. Tauzin, A., Gong, S.Y., Chatterjee, D., Ding, S., Painter, M.M., Goel, R.R., Beaudoin-Bussières, G., Marchitto, L., Boutin, M., Laumaea, A., et al. (2022). A boost with SARS-CoV-2 BNT162b2 mRNA vaccine elicits strong humoral responses independently of the interval between the first two doses. Cell Rep. 41. 10.1016/j.celrep.2022.111554.

17. Kurhade, C., Zou, J., Xia, H., Cai, H., Yang, Q., Cutler, M., Cooper, D., Muik, A., Jansen, K.U., Xie, X., et al. (2022). Neutralization of Omicron BA.1, BA.2, and BA.3 SARS-CoV-2 by 3 doses of BNT162b2 vaccine. Nat. Commun. 13, 3602. 10.1038/s41467-022-30681-1.

18. Kitchin, D., Richardson, S.I., van der Mescht, M.A., Motlou, T., Mzindle, N., Moyo-Gwete, T., Makhado, Z., Ayres, F., Manamela, N.P., Spencer, H., et al. (2022). Ad26.COV2.S breakthrough infections induce high titers of neutralizing antibodies against Omicron and other SARS-CoV-2 variants of concern. Cell Rep. Med. 3, 100535. 10.1016/j.xcrm.2022.100535.

19. Miyamoto, S., Arashiro, T., Adachi, Y., Moriyama, S., Kinoshita, H., Kanno, T., Saito, S., Katano, H., Iida, S., Ainai, A., et al. (2022). Vaccination-infection interval determines cross-neutralization potency to SARS-CoV-2 Omicron after breakthrough infection by other variants. Med, S2666634022000897. 10.1016/j.medj.2022.02.006.

20. Qu, P., Faraone, J., Evans, J.P., Zou, X., Zheng, Y.-M., Carlin, C., Bednash, J.S., Lozanski, G., Mallampalli, R.K., Saif, L.J., et al. (2022). Neutralization of the SARS-CoV-2 Omicron BA.4/5 and BA.2.12.1 Subvariants. N. Engl. J. Med. 386, 2526–2528. 10.1056/NEJMc2206725.

21. Tuekprakhon, A., Nutalai, R., Dijokaite-Guraliuc, A., Zhou, D., Ginn, H.M., Selvaraj, M., Liu, C., Mentzer, A.J., Supasa, P., Duyvesteyn, H.M.E., et al. (2022). Antibody escape of SARS-CoV-2 Omicron BA.4 and BA.5 from vaccine and BA.1 serum. Cell 185, 2422-2433.e13. 10.1016/j.cell.2022.06.005.

22. Wang, Q., Guo, Y., Iketani, S., Nair, M.S., Li, Z., Mohri, H., Wang, M., Yu, J., Bowen, A.D., Chang, J.Y., et al. (2022). Antibody evasion by SARS-CoV-2 Omicron subvariants BA.2.12.1, BA.4, & BA.5. Nature, s1–3. 10.1038/s41586-022-05053-w.

23. Shen, X., Chalkias, S., Feng, J., Chen, X., Zhou, H., Marshall, J.-C., Girard, B., Tomassini, J.E., Aunins, A., Das, R., et al. (2022). Neutralization of SARS-CoV-2 Omicron BA.2.75 after mRNA-1273 Vaccination. N. Engl. J. Med. 387, 1234–1236. 10.1056/NEJMc2210648.

24. Sheward, D.J., Kim, C., Fischbach, J., Sato, K., Muschiol, S., Ehling, R.A., Björkström, N.K., Hedestam, G.B.K., Reddy, S.T., Albert, J., et al. (2022). Omicron sublineage BA.2.75.2 exhibits extensive escape from neutralising antibodies. Lancet Infect. Dis. 0. 10.1016/S1473-3099(22)00663-6.

25. Arora, P., Kempf, A., Nehlmeier, I., Schulz, S.R., Jäck, H.-M., Pöhlmann, S., and Hoffmann, M. (2022). Omicron sublineage BQ.1.1 resistance to monoclonal antibodies. Lancet Infect. Dis. 0. 10.1016/S1473-3099(22)00733-2.

26. Kurhade, C., Zou, J., Xia, H., Liu, M., Chang, H.C., Ren, P., Xie, X., and Shi, P.-Y. (2022). Low neutralization of SARS-CoV-2 Omicron BA.2.75.2, BQ.1.1, and XBB.1 by 4 doses of parental mRNA vaccine or a BA.5-bivalent booster. 2022.10.31.514580. 10.1101/2022.10.31.514580.

27. Tauzin, A., Nayrac, M., Benlarbi, M., Gong, S.Y., Gasser, R., Beaudoin-Bussières, G., Brassard, N., Laumaea, A., Vézina, D., Prévost, J., et al. (2021). A single dose of the SARS-CoV-2 vaccine BNT162b2 elicits Fc-mediated antibody effector functions and T cell responses. Cell Host Microbe 0. 10.1016/j.chom.2021.06.001.

28. Tauzin, A., Gong, S.Y., Beaudoin-Bussières, G., Vézina, D., Gasser, R., Nault, L., Marchitto, L., Benlarbi, M., Chatterjee, D., Nayrac, M., et al. (2022). Strong humoral immune responses against SARS-CoV-2 Spike after BNT162b2 mRNA vaccination with a 16-week interval between doses. Cell Host Microbe 30, 97-109.e5. 10.1016/j.chom.2021.12.004.

29. Prévost, J., Gasser, R., Beaudoin-Bussières, G., Richard, J., Duerr, R., Laumaea, A., Anand, S.P., Goyette, G., Benlarbi, M., Ding, S., et al. (2020). Cross-Sectional Evaluation of Humoral Responses against SARS-CoV-2 Spike. Cell Rep. Med. 1, 100126. 10.1016/j.xcrm.2020.100126.

30. Anand, S.P., Prévost, J., Nayrac, M., Beaudoin-Bussières, G., Benlarbi, M., Gasser, R., Brassard, N., Laumaea, A., Gong, S.Y., Bourassa, C., et al. (2021). Longitudinal analysis of humoral immunity against SARS-CoV-2 Spike in convalescent individuals up to eight months post-symptom onset. Cell Rep. Med., 100290. 10.1016/j.xcrm.2021.100290.

31. Tauzin, A., Gendron-Lepage, G., Nayrac, M., Anand, S.P., Bourassa, C., Medjahed, H., Goyette, G., Dubé, M., Bazin, R., Kaufmann, D.E., et al. (2022). Evolution of Anti-RBD IgG Avidity Following SARS-CoV-2 Infection. Viruses 14, 532. 10.3390/v14030532.

32. Nayrac, M., Dubé, M., Sannier, G., Nicolas, A., Marchitto, L., Tastet, O., Tauzin, A., Brassard, N., Lima-Barbosa, R., Beaudoin-Bussières, G., et al. (2022). Temporal associations of B and T cell immunity with robust vaccine responsiveness in a 16-week interval BNT162b2 regimen. Cell Rep. 0. 10.1016/j.celrep.2022.111013.

33. Li, W., Chen, Y., Prévost, J., Ullah, I., Lu, M., Gong, S.Y., Tauzin, A., Gasser, R., Vézina, D., Anand, S.P., et al. (2022). Structural basis and mode of action for two broadly neutralizing antibodies against SARS-CoV-2 emerging variants of concern. Cell Rep. 38, 110210. 10.1016/j.celrep.2021.110210.

34. Prévost, J., Richard, J., Gasser, R., Ding, S., Fage, C., Anand, S.P., Adam, D., Vergara, N.G., Tauzin, A., Benlarbi, M., et al. (2021). Impact of temperature on the affinity of SARS-CoV-2 Spike glycoprotein for host ACE2. J. Biol. Chem., 101151. 10.1016/j.jbc.2021.101151.

35. Ullah, I., Prévost, J., Ladinsky, M.S., Stone, H., Lu, M., Anand, S.P., Beaudoin-Bussières, G., Symmes, K., Benlarbi, M., Ding, S., et al. (2021). Live imaging of SARS-CoV-2 infection in mice reveals that neutralizing antibodies require Fc function for optimal efficacy. Immunity, S1074-7613(21)00347-2. 10.1016/j.immuni.2021.08.015.

36. Gruell, H., Vanshylla, K., Tober-Lau, P., Hillus, D., Schommers, P., Lehmann, C., Kurth, F., Sander, L.E., and Klein, F. (2022). mRNA booster immunization elicits potent neutralizing serum activity against the SARS-CoV-2 Omicron variant. Nat. Med., 1–4. 10.1038/s41591-021-01676-0.

37. Garrett, N., Tapley, A., Andriesen, J., Seocharan, I., Fisher, L.H., Bunts, L., Espy, N., Wallis, C.L., Randhawa, A.K., Ketter, N., et al. (2022). High Rate of Asymptomatic Carriage Associated with Variant Strain Omicron. medRxiv, 2021.12.20.21268130. 10.1101/2021.12.20.21268130.

38. Sun, K., Tempia, S., Kleynhans, J., von Gottberg, A., McMorrow, M.L., Wolter, N., Bhiman, J.N., Moyes, J., du Plessis, M., Carrim, M., et al. (2022). SARS-CoV-2 transmission, persistence of immunity, and estimates of Omicron’s impact in South African population cohorts. Sci. Transl. Med. 0, eabo7081. 10.1126/scitranslmed.abo7081.

39. Altarawneh, H.N., Chemaitelly, H., Ayoub, H.H., Hasan, M.R., Coyle, P., Yassine, H.M., Al-Khatib, H.A., Benslimane, F.M., Al-Kanaani, Z., Al-Kuwari, E., et al. (2022). Protection of SARS-CoV-2 natural infection against reinfection with the Omicron BA.4 or BA.5 subvariants. 2022.07.11.22277448. 10.1101/2022.07.11.22277448.

40. Carazo, S., Skowronski, D.M., Brisson, M., Sauvageau, C., Brousseau, N., Gilca, R., Ouakki, M., Barkati, S., Fafard, J., Talbot, D., et al. (2022). Protection against Omicron re-infection conferred by prior heterologous SARS-CoV-2 infection, with and without mRNA vaccination. 2022.04.29.22274455. 10.1101/2022.04.29.22274455.

41. Goel, R.R., Apostolidis, S.A., Painter, M.M., Mathew, D., Pattekar, A., Kuthuru, O., Gouma, S., Hicks, P., Meng, W., Rosenfeld, A.M., et al. (2021). Distinct antibody and memory B cell responses in SARS-CoV-2 naïve and recovered individuals following mRNA vaccination. Sci. Immunol. 6. 10.1126/sciimmunol.abi6950.

42. Valcourt, E.J., Manguiat, K., Robinson, A., Lin, Y.-C., Abe, K.T., Mubareka, S., Shigayeva, A., Zhong, Z., Girardin, R.C., DuPuis, A., et al. (2021). Evaluating Humoral Immunity against SARS-CoV-2: Validation of a Plaque-Reduction Neutralization Test and a Multilaboratory Comparison of Conventional and Surrogate Neutralization Assays. Microbiol. Spectr. 9, e0088621. 10.1128/Spectrum.00886-21.

43. Sun, H., Xu, J., Zhang, G., Han, J., Hao, M., Chen, Z., Fang, T., Chi, X., and Yu, C. (2022). Developing Pseudovirus-Based Neutralization Assay against Omicron-Included SARS-CoV-2 Variants. Viruses 14, 1332. 10.3390/v14061332.

44. Wang, Z., Muecksch, F., Muenn, F., Cho, A., Zong, S., Raspe, R., Ramos, V., Johnson, B., Ben Tanfous, T., DaSilva, J., et al. (2022). Humoral immunity to SARS-CoV-2 elicited by combination COVID-19 vaccination regimens. J. Exp. Med. 219, e20220826. 10.1084/jem.20220826.

45. Schmidt, F., Muecksch, F., Weisblum, Y., Da Silva, J., Bednarski, E., Cho, A., Wang, Z., Gaebler, C., Caskey, M., Nussenzweig, M.C., et al. (2022). Plasma Neutralization of the SARS-CoV-2 Omicron Variant. N. Engl. J. Med. 386, 599–601. 10.1056/NEJMc2119641.

46. Robbiani, D.F., Gaebler, C., Muecksch, F., Lorenzi, J.C.C., Wang, Z., Cho, A., Agudelo, M., Barnes, C.O., Gazumyan, A., Finkin, S., et al. (2020). Convergent antibody responses to SARS-CoV-2 in convalescent individuals. Nature 584, 437–442. 10.1038/s41586-020-2456-9.

47. Jennewein, M.F., MacCamy, A.J., Akins, N.R., Feng, J., Homad, L.J., Hurlburt, N.K., Seydoux, E., Wan, Y.-H., Stuart, A.B., Edara, V.V., et al. (2021). Isolation and characterization of cross-neutralizing coronavirus antibodies from COVID-19+ subjects. Cell Rep. 36, 109353. 10.1016/j.celrep.2021.109353.

48. Gong, S.Y., Chatterjee, D., Richard, J., Prévost, J., Tauzin, A., Gasser, R., Bo, Y., Vézina, D., Goyette, G., Gendron-Lepage, G., et al. (2021). Contribution of single mutations to selected SARS-CoV-2 emerging variants spike antigenicity. Virology 563, 134–145. 10.1016/j.virol.2021.09.001.

49. Tauzin, A., Gong, S.Y., Beaudoin-Bussières, G., Vézina, D., Gasser, R., Nault, L., Marchitto, L., Benlarbi, M., Chatterjee, D., Nayrac, M., et al. (2022). Strong humoral immune responses against SARS-CoV-2 Spike after BNT162b2 mRNA vaccination with a 16-week interval between doses. Cell Host Microbe 30, 97-109.e5. 10.1016/j.chom.2021.12.004.

50. Beaudoin-Bussières, G., Laumaea, A., Anand, S.P., Prévost, J., Gasser, R., Goyette, G., Medjahed, H., Perreault, J., Tremblay, T., Lewin, A., et al. (2020). Decline of Humoral Responses against SARS-CoV-2 Spike in Convalescent Individuals. mBio 11. 10.1128/mBio.02590-20.

51. Ding, S., Laumaea, A., Benlarbi, M., Beaudoin-Bussières, G., Gasser, R., Medjahed, H., Pancera, M., Stamatatos, L., McGuire, A.T., Bazin, R., et al. (2020). Antibody Binding to SARS-CoV-2 S Glycoprotein Correlates with but Does Not Predict Neutralization. Viruses 12, 1214. 10.3390/v12111214.

